# Community Organizing for Indigenous People in the Philippines: A Proposed Approach

**DOI:** 10.1101/2021.06.24.21259509

**Authors:** Jailah Bamba, Cristela Candelario, Rosarie Gabuya, Lhearnie Manongdo

**Affiliations:** College of Public Health, University of the Philippines – Manila

**Keywords:** community organizing, community participation, indigenous people, geographically isolated and disadvantaged areas, indigenous Filipinos

## Abstract

Cognizant of the special needs of indigenous people in the Philippines, the Republic Act No. 8371 of 1997 was established to promote and protect their rights. Over the years, a number of community organizing efforts for the improvement of these communities were conducted by stakeholders from the private and public sectors. However, resistance has been reported due to poor understanding and integration of these indigenous populations’ varied cultures and traditions. This study aims to describe the predominant principles and frameworks used for community organizing among indigenous people. Specifically, it seeks to propose a community organizing approach that is culturally sensitive and appropriate for indigenous communities in geographically isolated and disadvantaged areas in the Philippines. A systematic review was conducted on four databases (PubMed, ScienceDirect, ResearchGate, Google Scholar) by four independent researchers. Inclusion criteria involved studies about community organizing protocols in the Philippines, published in peer-reviewed journals from 2010-2020, and written in the English language. Assessment of the quality of included studies was done using the Critical Appraisal Skills Program (CASP) checklist, and narrative synthesis was employed to summarize and report the findings. Thirteen studies met our inclusion criteria out of a total of fifty-five articles searched. Based on the evidence, our proposed approach builds on Groundwork, Indigenous Capacity Building, Community Participation and Ownership, Mobilization, and Sustainability. We highlight the emphasis of harnessing indigenous knowledge and Participatory Monitoring and Evaluation to involve them in all steps of the planning and decision-making processes. Furthermore, we distill tools and methodologies that could strengthen and precipitate successful community organizing endeavors.

## 1.0 INTRODUCTION

### 1.1 Background of the Study

The Philippines is a culturally diverse country with various ethnolinguistic groups that denote genealogical, paternal as well as maternal lineage to any of the country’s group of native population^1^. According to the 2015 Population Census, Indigenous People (IP) in the Philippines constitute 10%-20% of the national population of 100,981,437. The estimated 14-17 million IPs belong to 110 ethno-linguistic groups, which are mainly concentrated in Northern Luzon (Cordillera Administrative Region, 33%) and Mindanao (61%), with some groups in the Visayas area ^1^.

Republic Act 8371, known as “The Indigenous Peoples’ Rights Act (IPRA) of 1997”, states that Indigenous Cultural Communities/Indigenous People are:

> *“a group of people or homogenous societies identified by self-ascription and ascription by others, who have continuously lived as an organized community on communally bounded and defined territory, and who have, under claims of ownership since time immemorial, occupied, possessed and utilized such territories, sharing common bonds of language, customs, traditions, and other distinctive cultural traits, or who have, through resistance to political, social and cultural inroads of colonization, nonindigenous religions, and cultures became historically differentiated from the majority of Filipinos*.*”*

In the Philippines, the IP communities remain among the poorest and most disadvantaged peoples. Because they have retained their traditional pre-colonial culture and practices, they were subjected to discrimination and few opportunities for major economic activities, education, or political participation. As a result, they have been resistant to development and information, thus have been driven to geographically isolated disadvantaged areas (GIDAs) with no adequate and accessible basic services.

To recognize this diversity, the Philippine Constitution signed Republic Act No. 8371 of 1997, which seeks to identify, promote, and protect the rights of the IPs. These include the Right to Ancestral Domain and Lands; Right to Self-Governance and Empowerment; Social Justice and Human Rights; and the Right to Cultural Integrity ^3^, which is in line with the United Nations Declaration on the Rights of Indigenous Peoples in 2007.

Despite the international and national recognition of the inherent rights of indigenous peoples, IP communities were subjected to historical discrimination and marginalization from political processes and economic benefits. The Report on the State of the World of Indigenous Peoples, issued by the United Nations Permanent Forum on Indigenous Issues in January 2010, revealed that IPs’ traditional livelihoods were threatened by extractive industries or substantial development projects. Still, they continued to be over-represented among the poor, the illiterate, and the unemployed. While they constitute approximately 5 percent of the world’s population, IPs make up 15 percent of the world’s poor, comprise about one-third of the world’s 900 million destitute rural people, continuously suffer disproportionately in areas like health, education, and human rights, and regularly face systemic discrimination and exclusion ^3^.

In the Philippines, the National Commission on Indigenous People (NCIP) and Department of Social Welfare and Development were mandated to implement programs, projects, and provide services through engaging the indigenous people in a meaningful development process where there is full recognition of their capacity to strengthen their own economic, social, and political systems ^4^. As a result, the IP sector has a broad spectrum of active support groups and organizations from government, academe, non-government organizations, international groups, and churches. In addition, the enactment of IPRA paved the way for the growth of IP support groups that provide assistance on policy advocacy, education, community development, and poverty alleviation programs.

However, despite the growing number of NGOs and programs for IP communities, conflicts among the community members continuously rise. In an attempt by the State to enforce developmental projects, there was growing resistance from IPs due to their own indigenous governance, which struggles to preserve their own customary laws and traditions. There were also instances wherein NGOs with little or no exposure to IPs cultures have generated conflicts because of insufficient program analysis. In addition, pressure from funding donors who have tight project schedules and are pushed to produce outcomes has resulted in shortcuts, thus marginalizing critical community processes ^5^.

This review aims to describe studies that have meaningful participation of IP communities and explore ways to tailor the community organizing principles to be more culturally sensitive in improving IPs’ access to essential social services such as health, nutrition, sanitation, and formal and non-formal education. Culture-sensitive facilitation is defined as awareness and acceptance of cultural differences on the part of the facilitator that allow them to engage with indigenous people in a manner that is appropriate and responsive to their customs, traditions, values, and beliefs ^4^. This regards IP communities not just mere passive recipients or beneficiaries but as active partners in the program. The community organizing principles presented may help community organizers, NGOs, and even government programs to provide a more sustainable implementation of the activities. The study focuses on the IPs in geographically isolated and disadvantaged areas, particularly those with: no or limited opportunities for development, no or limited access to social services, no access road or hard to reach areas, and insufficiency of food security.

Based on a thorough review of research evidence, the proposed community organizing approach can be applied for the implementation of health programs and interventions that are culturally sensitive and responsive. This would support the paradigm shift in participation practices that value community input while minimizing risks of unintended harms and consequences for indigenous communities.

### 1.2. Literature Review

Community organizing is the process by which the people organize themselves to ‘take charge’ of their situation and thus develop a sense of being a community together ^6^. It is one of the strategies that empower disadvantaged communities to promote social change and behavior. Several studies were conducted to extend the current knowledge base of community organizing to build formal community organizing practice theory grounded in the literature and expertise of skilled community organizers ^6^. These frameworks and protocols were used by international and local organizations in implementing their goals and helped develop communities that are challenged with unjust systems and policies.

#### Community Organizing in the International Community

The importance of community organizing was seen in a study done in the United States that promoted health and services in disenfranchised populations that cannot afford medical care in an affluent community ^7^. The study by Bezboruah showed that community organizing among nonprofit organizations, civic bodies, citizens, and other grassroots organizations is effective in providing direct services and influencing public policies through their advocacy activities. Another notable finding in the study is the role of a facilitator organization that will act as a mediator in order to improve collaboration and coordination among stakeholders. These facilitators can persistently uphold the community organizing goals of building trust and generating commitment from community members through formal and informal mechanisms ^7^.

On the other hand, community organizing can also rally mass momentum for social transformation by changing the policy, challenging public resources allocation, and transforming realities on the ground. This was also observed in different states such as California ^8^, Detroit ^9^, Mississippi Delta ^10^, which all led to successful movements and campaigns across race, faith, and gender. Generally, participatory action strategies were mostly applied in community organizing in order to empower participants and transfer the accountability to the community while at the same time valuing participants’ voices. Another trend observed in community organizing internationally is that organizers played the role of an interpreter that shared program ownership rather than mere implementers of the program.

#### Community Organizing in the Philippines

In the Philippines, community organizing was used by social development workers in empowering people’s organizations to address poverty and social inequality ^11^. As early as 1985, several events were held to assess community organizing practices and future development of the strategy to enhance the capabilities and resources of the community over the past years. Among these activities include Integration, Social Investigation, and Issue Identification and Analysis. In these steps, there was a need for an organizer to immerse himself with the local community in order to systematically learn and analyze the structures and forces within the community. Learning the issues and problems in the community by experience rather than in secondary data helped the development workers define and prioritize community problems. Since community organizing is an enabling process, the People’s Organization usually takes over the organizer’s role when the indicators such as high level of socio-political awareness, sustained membership participation, and presence of trained community leaders were significantly met ^11^.

Community organizing was also observed in rural communities in the Philippines. A study done in San Jose and Kagawasan, wherein a large majority of the communities were poor and situated in mountainous areas of Manobo, showed that community participation was affected not just by individual decisions but also by constraints on actions imposed by bureaucratic structures and restrictive institutional arrangements ^12^. Thus, community organizing was used to eliminate these barriers, thereby helping the community members grow in efficacy, self-awareness, and confidence in their future. The community was transformed from traditionally having a passive stance when faced with authorities, to having a collective power and transforming them into a dynamic and thinking group. The study applied a community organizing “bottom-up” strategy approach wherein members organized themselves because they were aware of the grassroots issues of the community. Furthermore, the tactics used in community organizing were sustained even when the community organizers left the place.

#### Community Organizing in Indigenous People

Indigenous peoples in the Philippines have retained much of their traditional pre-colonial culture, and often situate themselves in geographically isolated disadvantaged areas of the country. There is a great variety of social organization and cultural expression among these people, and thus penetrating into these communities is difficult during implementation of programs and community organizing efforts.

In order to fulfill its mandate to develop, administer and implement social services to the disadvantaged and marginalized sectors, the Department of Social Welfare and Development (DSWD) executed Memorandum Circular OI, Series of 2009 known as Indigenous Peoples Policy Framework (IPPF). This is in conjunction with the Indigenous Peoples Rights Act or (IPRA Law) and UN Declaration on the Rights of the Indigenous Peoples. The IPPF was formulated to serve as a “declaration of policies and standard procedures in developing, funding and implementing programs, projects and services for indigenous peoples ^13^”. The following strategies were included in the framework: 1) Situational Analysis/Project Identification 2) Project Planning 3) Project Appraisal 4) Project Implementation, Operation, and Management 5) Progress Monitoring and Project Evaluation 6) Program Documentation 7) Project Localization/Sustainability. The following activities aimed to identify the programs that are acceptable and inclusive/responsive to their current and emerging needs. The framework focused on the actual consultation in order to determine the projects that are socially acceptable to their cultures, which was also in line with the community organizing strategy used by the international community.

A decade has passed but this framework is still being adopted by the DSWD in the implementation of programs and reform agenda to ensure full and sustainable IP participation and empowerment. Recently, the Indigenous People Framework was used for the Pilot Implementation of the Modified Conditional Cash Transfer Program for IPs in geographically isolated disadvantaged areas. Community organizing was mentioned as one of the program strategies by the DSWD as this facilitates the development of a community-based system that allows the IP Communities to participate in the planning and operations of the program ^14^. One goal of the community organizing process in this program was to have a collective consent of the community to proceed with the program prior to implementation. This collective consent was referred to as “Memorandum of Understanding” and is a prerequisite unique in the program implementation for IPs due to their rights as mentioned in the IPRA Law.

One major factor contributing to the IPs disadvantaged position is the lack of access to culture-responsive basic education. Thus, to support the DSWD’s framework, the Department of Education (DepEd) also adopted a National Indigenous Peoples Education Policy Framework which aimed to promote shared accountability, continuous dialogue, engagement, and partnership among government, IP communities, civil society, and other education stakeholders. Identical to the framework used by DSWD, the Indigenous Peoples Education (IPEd) Program is DepEd’s response to the right of indigenous peoples (IP) to basic education that is responsive to their context, respects their identities, promotes the value of their indigenous knowledge, skills, and other aspects of their cultural heritage ^15^. This program used a “rights-based approach” which focuses on the importance of the principles of participation, inclusion, and empowerment. This shows that across different institutions of the government, the Indigenous People Framework was used in order to have flexible, demand-driven, and evidence-based programs.

### 1.3 Theoretical Perspective

#### Principles of Community Organizing

The establishment of Indigenous People Policy Framework is also governed by some principles. Community Organizing as a process and a method is based on certain basic principles, which serve as guidelines to sound or effective practice^16^. Several literatures have discussed the different principles of community organizing. Some define the lines according to which a community organizing perspective must operate and these principles might also be applied among indigenous communities ^17^:

- Community organizing involves consciousness-raising through experiential learning. Central to the community organizing process is the development of awareness and motivation among the people to act upon their problems. As conscientization is achieved through practice, community organizing therefore emphasizes learning that emerges from concrete actions;
- Community organizing is participatory and mass-based. It involves the whole community in organizing experiences and is primarily directed towards and biased in favor of the poor. People must be mobilized to participate fully in all aspects of community organizing activities;
- Community organizing is based on democratic leadership. It is group-centered, not leader-oriented. Leaders emerge and are tested through concrete action, not externally appointed or selected. Hence, leaders are accountable to the people at all times;
- Community organization must work towards people’s empowerment so that they may liberate themselves from their oppression ^18^.

#### Approaches and Models of Community Organizing

Various authors coined community organizing approaches based on literature. Three approaches were identified which depend on the objective of the group, namely: 1) specific content objective, wherein “an individual, an agency, or an organization becomes concerned about a needed reform in the community and launches a program to secure this reform”, 2) general content objective, “group, association, or a council focused on the coordinated and orderly development of services in a particular area of interest”, and 3) process objective, where “the group aims to initiate and nourish a process in which all the people of a community are involved, through their representatives, in identifying and taking action about their own problems.” In this approach, the aim is to increase motivation, responsibility, and skill in recognizing and securing reforms the community considers desirable; and development of community integration and capacity to function as a unit with respect to community problems ^19^.

Other approaches used are project and political action. Project approach “attempts to organize communities around certain projects that aim for community self-reliance” while political action approach “focuses on collective action in which the community makes known its grievances and its demands to relevant authorities or to the public.”^16^

In practice, many community development workers employ a range of techniques, approaches, and models. Rothman enumerates three models, namely: *locality development, social planning, and social action* ^20^. *Locality development model* holds that community changes can be pursued most effectively by widely involving the local people in determining and achieving goals. It is used when populations are homogeneous or when consensus exists among various community subparts and interests. *Social planning*, on the other hand, necessitates the services of experts in effecting planned change processes, especially in solving social problems. It is adopted when community problems are fairly routine and can be solved through the application of factual information. Lastly, *Social action*, which is premised on the belief that there are disadvantaged segments in society that need to be organized to enable them to voice out their demands for social justice or democracy. It is suitable when community subgroups are hostile and interests are not reconcilable through usual discussion methods.

However, a study was conducted to explore culture and cultural competence, as it relates to community organizing as a modification to Rothman’s community organizing analysis framework, that will allow it to be used to identify and be responsive to the dynamics of culture in community practice ^21^. A number of community organizing models have emerged that consider culture as an influencing factor. The depth of culture used by these models varies, with some focused on building cross-cultural understanding while others offer organizing models centered in the world-view of specific ethnic groups. One of which is a model for intercultural organizing that mirrors cultural competence concepts. The model consists of dynamics, including understanding one’s own social location and cultural perspectives, in order to develop cultural sensitivity ^22^. It also discusses expanding one’s knowledge about other cultures and suggests that recognizing and building on community strengths is a core component of organizing with diverse communities. The model specifically emphasizes the need for empowerment efforts, along with recognizing and promoting indigenous leadership as well as the use of collaborative partnerships with community members, wherein power differences inherent in the helping relationship are minimized by the social worker assuming the dual role of facilitator and learner. Finally, the model emphasizes being able to recognize, understand, and diffuse inter- and intra-group conflict.

Some studies have discussed other models applied specifically to indigenous communities. One of which is the *village-based indigenous planning (VIP) model* based from the case study conducted among Mt. Apo Bagobo-Manobo sister tribes in the Philippines ^23^. This model covers a broad development viewpoint based on survey results and is supported by the holistic worldviews of indigenous people in general. Its underlying perspective for development is adaptation and holism. It was then concluded that the VIP model can supplement the existing Ancestral Domain Sustainable Development and Protection Plan (ADSDPP) process because it can address some weaknesses of the government framework. On the other hand, an eight-step approach for engaging communities was proposed in Africa^24^. The said approach was useful in the prevention and control of Ebola infection and was aligned with the Community, Assets, Responsiveness and Evaluation (CARE) Model, which was being developed for frontline infectious disease control practitioners. This model offers guidance for the development, implementation, and evaluation of community-engaged responsive and culturally congruent control efforts towards prevention, treatment, containment, and self-care of Ebola and other VHFs. It was stated that aside from rigorous adherence to standard infection prevention and control (IPC) precautions and safety standards, it is more successful if it is combined with the local community’s knowledge and experiences. The proposed eight-step model starts from entering communities with cultural humility, through reciprocal learning and trust, multi-method communication, development of the joint protocol, to assessing progress and outcomes and building for sustainability.

#### Theoretical framework of community organizing

The theoretical framework provides that community organizing is a process that aims to achieve a people-centered development for resource-poor communities characterized by passivity, dependency, and powerlessness ^16^.

As shown in Figure 1, this kind of development is only possible if people are conscientious, socially-transformed, self-reliant, participative, and empowered. These desired characteristics can be achieved through the community organizing process.

**Figure 1.**
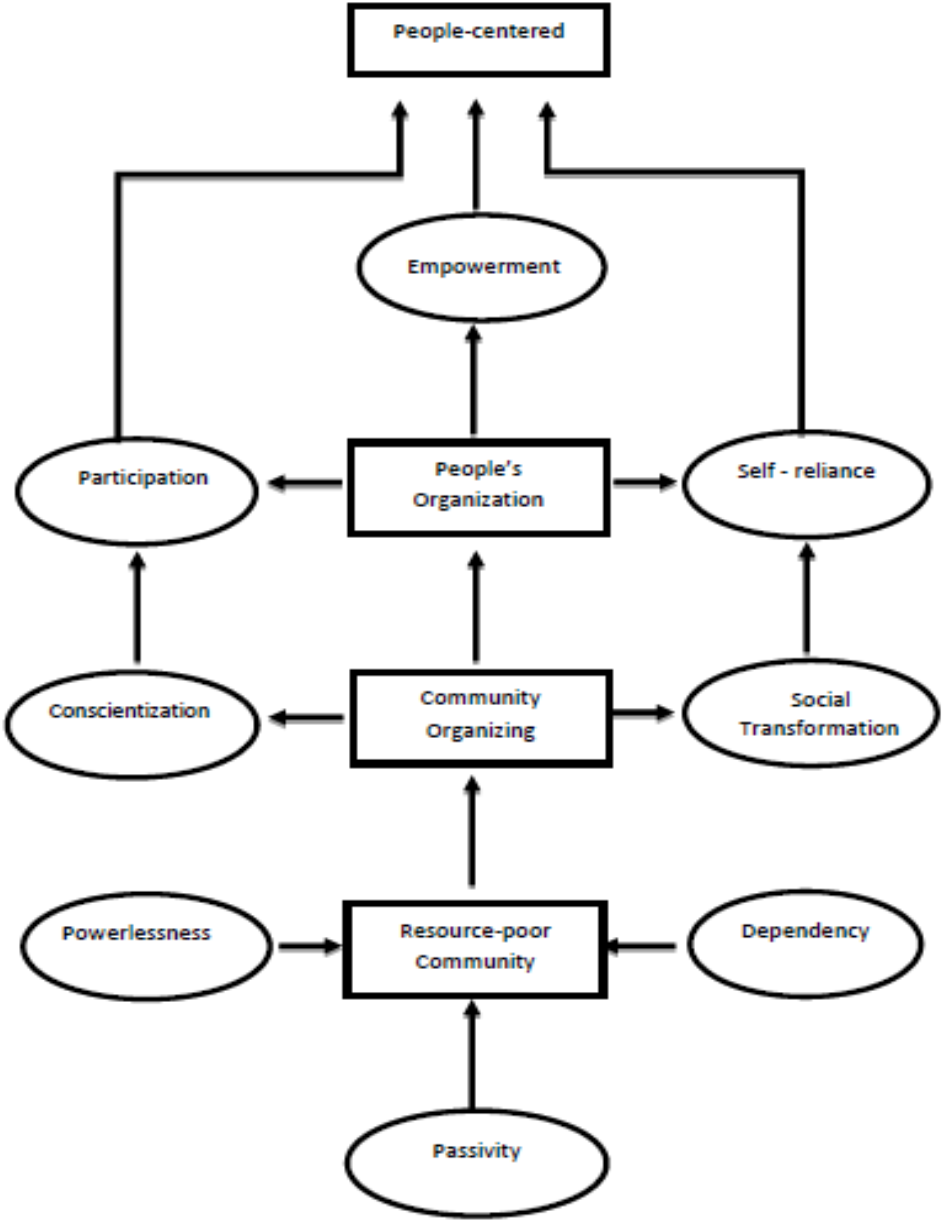
Theoretical Framework of Community Organizing

#### Critical Factors in Successful Indigenous Community-Managed Programs

There are several factors that can contribute to the success of indigenous community-managed programs^25^. The said study outlines the reasons for success in indigenous organizations and this include the following:

##### Facilitating community ownership and control

Community ownership ensures authority and autonomy in over all aspects of the project/program where it builds commitment and enthusiasm in all people involved. Evidence from qualitative studies suggests that community ownership and control can be embedded in community-managed programs in various ways which includes (but are not limited to): *1) forming local indigenous management bodies* - where the members were either totally or majority Indigenous community members. These bodies play an important role in making major decisions relating to the program. They also serve as conduits for community perspectives, which also coordinates with government agencies and sets strategic directions over projects to achieve its goals and objectives; *2) formal agreement with partner organizations* – this include written agreements to provide clarity with partner organizations. It also establishes the indigenous organization’s strategic vision and any mutual agreements over particular matters.

##### Embedding culture

Maintaining the indigenous culture is also essential in the success of the programs. These programs should be built around positive cultural perceptions. Moreover, it was also mentioned that prioritizing the indigenous worldview is an important aspect of embedding culture wherein it should be relationally and holistically based on community and family obligations rather than the individual.

##### Employing local indigenous staff

Employing local indigenous staff can help in effectively communicating the program in appropriate language and in a way that is also suited with the local social and cultural values. The local indigenous staff has also an important leadership role for community members.

##### Implementing good governance and establishing trusting partnerships

Building good governance is essential in building sustainable development in communities. An important aspect of good governance in Indigenous communities is achieving a legitimate cultural fit. While complex in practice, a cultural fit in the context of governance involves a balance between organizational governance standards and community traditions and values. Moreover, having strong and trusting relationships with partner organizations is also a key success factor in effective indigenous-managed programs.

##### Using community development approaches

Often, successful indigenous community-managed programs mirror the key community development principles of bottom-up development, empowerment, community ownership and decision making, and prioritize IP’s strong connection to land, family and culture. The approaches and strategies should be appropriate to the type of indigenous communities, and recognizing and adjusting the practices to local differences is therefore essential.

### 1.4 Research Objective

The study aimed to describe the predominant principles, frameworks, and appropriate methodologies used for community organizing among Indigenous People. Specifically, it also aimed to propose a community organizing approach that is culturally sensitive and appropriate for indigenous communities in geographically isolated and disadvantaged areas in the country.

## 2.0 METHODS

### 2.1 Study Design

The study utilized a systematic review design to describe the approaches to community organizing for IPs in GIDAs in the Philippines. A systematic review enables the researchers to start with a well-defined research question, find all existing evidence in an unbiased, transparent, and reproducible way, and provide recommendations based on knowledge gaps^26^. Systematic reviews seek to answer clearly formulated questions by using rigorous, explicit protocols to identify, select, and appraise relevant research studies; and to collect and analyze data from the selected studies.^27^

### 2.2 Search Strategy and Data Sources

The researchers independently conducted elementary analysis on four online databases namely: ResearchGate, PubMed, ScienceDirect, and Google Scholar to identify relevant and published studies. An elementary search on Titles and Abstracts was conducted in May 11, 2021 which identified a range of available evidence on the community organizing frameworks or protocols implemented for IPs in local and international settings. Further search was conducted on May 18, 2021 to update the results. Keywords used were: Community organizing and Philippines, Community participation and Philippines, Community engagement, Community organizing for Indigenous People, Indigenous people and planning, Indigenous people in Asia and Philippines, Indigenous Planning Framework, Community Mobilization, Community Participation, Rural development, Collective Action. In addition, the researchers reviewed the selected articles’ references in order to identify additional studies or reports not retrieved by the preliminary searches (reference by reference). Upon completion of the data collection and preliminary screening, the researchers convened and reviewed the articles individually.

### 2.3 Study Selection

This systematic review utilized the PRISMA 2020 (Preferred Reporting Items for Systematic Reviews and Meta-Analyses) guidelines. PRISMA is an evidence-based minimum set of items for reporting in systematic reviews and meta-analyses. This is useful for critical appraisal of published systematic reviews.^26^ The PRISMA flow chart demonstrates the step-by-step process of identification and screening of potentially eligible studies and determines the final number of studies included for analysis based on inclusion and exclusion criteria. This consists of the 3 steps of sample selection, namely: 1) Identification, 2) Screening, 3) Included (see Figure 2).

**Figure 2.**
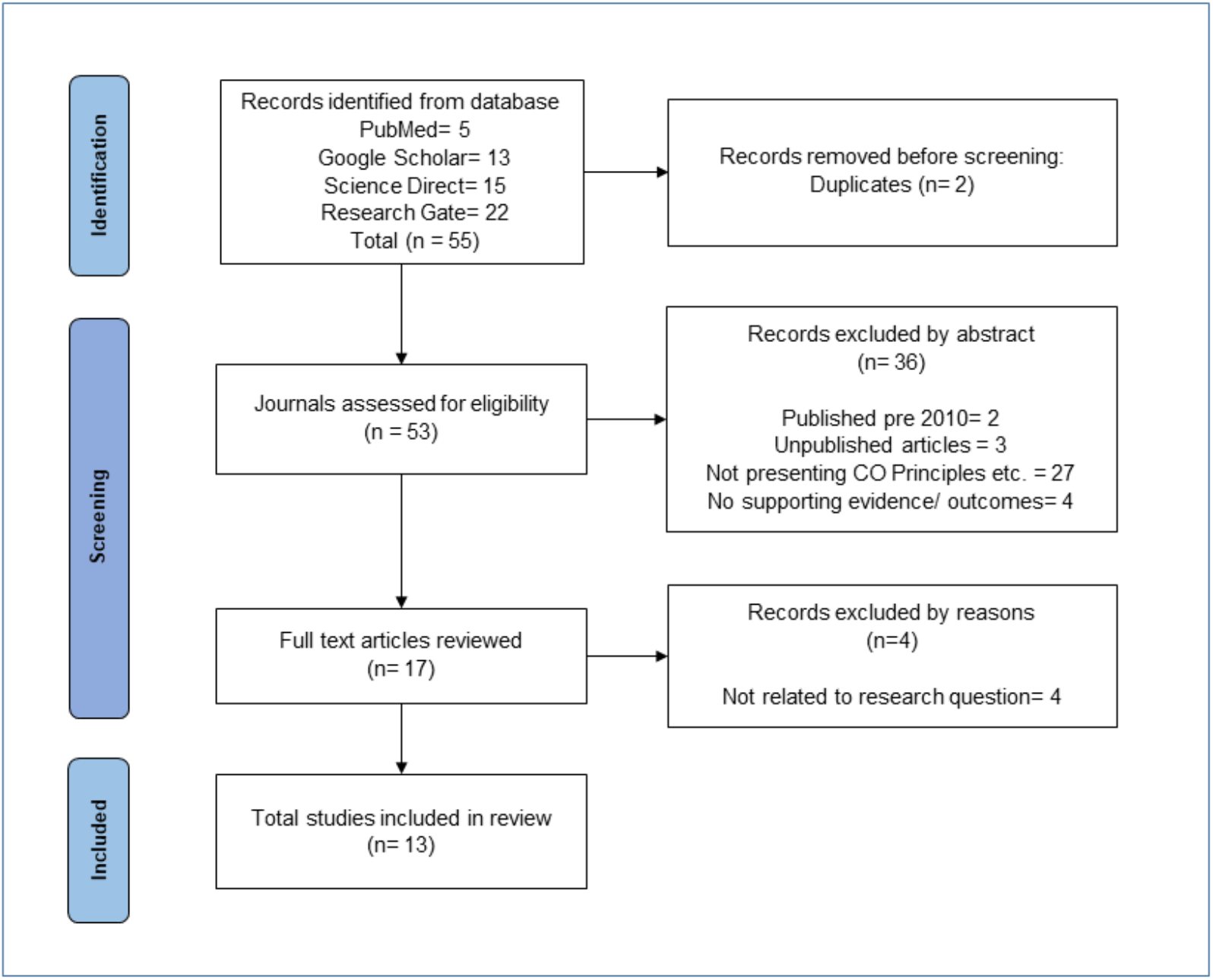
PRISMA flow diagram

After independently searching the aforementioned databases, a total of 55 journal articles returned from the keywords used. Data cleaning then commenced to remove duplicates leaving 53 articles to assess for eligibility. Using the inclusion criteria on the titles and abstracts, about 36 articles were removed as they did not fit with the parameters such as being published pre-2010, articles from non-peer reviewed journals, those that did not employ community organizing principles, and those without supporting evidence/outcomes. This resulted in 17 full-text articles that were screened and reviewed. These were further narrowed down to remove those that were not relevant to the research objective. A final count of 13 journal articles were included in the final review.

### 2.4 Eligibility Criteria

All studies searched from the identified databases that showed concepts of community development, organizing, and mobilization targeting the local communities such as Indigenous People and Geographically Isolated Disadvantaged Areas, were included in the preliminary analyses. The following inclusion and exclusion criteria were applied to identify the eligible articles to be reviewed.

### 2.5 Data Extraction

After the initial screening, full-text of the study was retrieved and was subjected to quality assessment. Data were extracted from all research papers that met the inclusion criteria. The following data were extracted and analyzed using the following criteria: first author, year of publication, title of the study, country, target population, intervention, comparison, community organizing principles used, outcome, and study design.

### 2.6 Quality Assessment

After extracting the papers which met the eligibility criteria, assessment of the quality of each paper was then carried out using the Critical Appraisal Skills Program (CASP) checklist.

The CASP is a template developed by a team of experts to guide people how to critically appraise different types of research evidence ^24^. This tool allowed the researchers to gauge the clarity of each article’s study objectives, the quality of the methodology, research design, data collection and analyses, ethical considerations, whether there was a clear statement of findings, and the overall value of the research.

### 2.7 Evidence Synthesis

In order to summarize and explain the findings of the multiple studies appraised, narrative synthesis was employed. A narrative synthesis is a form of story-telling wherein evidence is brought together in a systematic way so as to make sense of the data and answer a research question ^25^.

This was done by first developing a preliminary synthesis of the topic of the papers (using the PICOS parameters of Population, Intervention, Comparison, Outcome, and Study Design), exploring relationships within and between studies to determine patterns and trends, and assessing the robustness of the synthesis by considering the methodological quality of the papers being reviewed (such as the quality and quantity of the evidence base it is built on).

The researchers held frequent meetings to discuss the findings from the articles they independently assessed.

### 2.8 Ethical Considerations

Since this systematic review only worked on primary research studies and the researchers have no direct access to any of the participants, it is exempted from the Ethics Review board. However, a critical reflection upon the contextual position of the authors included in the review was still upheld. This is to ensure that the evaluation and interpretation of evidence from the selected studies were not refracted through the subjective lens of authors of individual studies^26^.

This was done using a couple of methods - first, by way of ethically considering the quality and relevance of evidence reported using evaluation criteria that were aligned with the overarching methodological orientation of our review. This is important so as to ethically reflect on any information that may be missing and how this may influence the report findings.

Second, through taking note of constructs in trustworthiness that are evident in scientifically sound qualitative studies (i.e. credibility, transferability, dependability, and confirmability) to avoid potential biases. And third, by means of recognizing the epistemological positioning of the original study to distill information that was most relevant for addressing the review’s purpose. All these factors helped shape the findings of this review.

## 3.0 SEARCH RESULTS

### 3.1 Quality of the Included Studies

The following section shows the results of the Critical Appraisal Skills Program (CASP) quality assessment of the included studies using the standardized checklist (see Appendix). Most of the included studies were able to pass the parameters set by the tool (*n = 10*) with three studies that were not able to fully satisfy one or more parameters of quality (see Table 2).

**Table 1.** Inclusion and Exclusion Criteria

| INCLUSION CRITERIA | EXCLUSION CRITERIA |
| --- | --- |
| Articles published from 2010 to 2020<br>Journals written in the English language<br>Target population included the Indigenous People or people in GIDAs<br>Published journal articles, manuals / guidebooks, annual reports, project reports (government and NGO issuances)<br>Culturally sensitive - All information from the specific articles should promote the rights of ICCs/IPs based from United Nations Declaration on the Rights of Indigenous Populations<br>Articles including community improvement but not limited to existing frameworks or models of community organizing, community mobilization, community engagement, and community participation | Review articles, opinion articles, news articles, unpublished research, manuscripts, (conference papers, pre-prints, etc.)<br>Studies not related to the research question<br>Papers presenting result without supporting evidence and results of study |

**Table 2.**
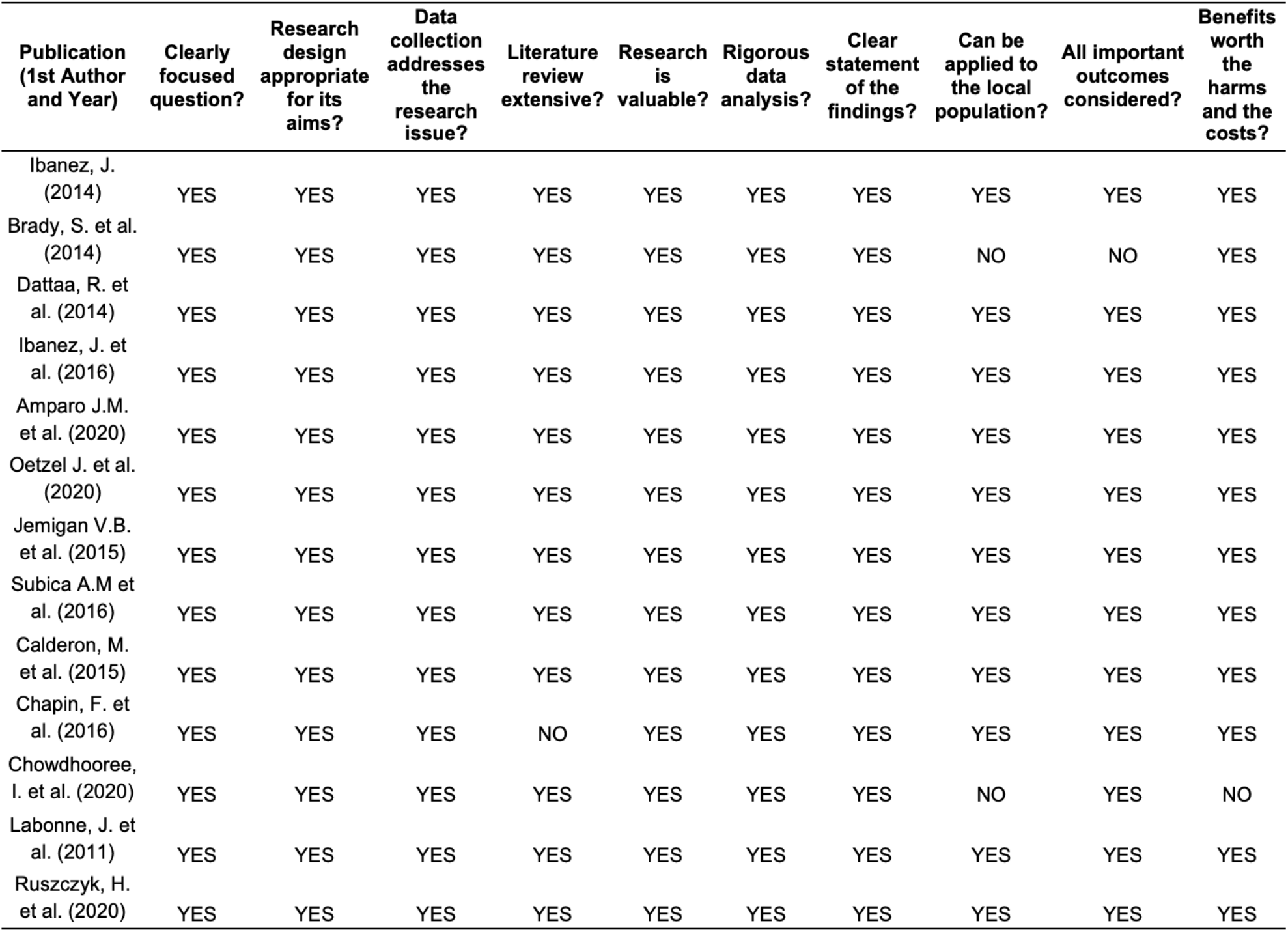
Critical Appraisal Skills Program (CASP) Quality Assessment of Included Studies

### 3.2 Characteristics of the Included Studies

From a total of 55 research articles searched using pertinent keywords, a total of 13 papers met our set inclusion and exclusion criteria after removing duplicates and thorough data cleaning and parsing. The included studies published in various international journals were relatively recent: 2011 (*n = 1)*, 2014 (*n = 3)*, 2015 (*n = 2)*, 2016 (*n = 3)*, 2020 (*n = 4)*. Five studies were carried out in the Philippines while eight studies were conducted abroad. Among the studies that were done internationally, five were done in high-income countries (e.g. New Zealand and the United States of America) and three were from lower-middle-income countries (e.g. Nepal and Bangladesh).

Based on the design of the study, three studies were experimental-exploratory, three were case studies, two were qualitative, and one each using survey, action research, cohort, mixed methods, and descriptive quantitative research designs. Most of the studies involved partnerships with nongovernmental organizations *(n = 11)* while some sought the help of academic institutions *(n = 2)*, especially where coordination with other organizations and training was required. Table 3 summarizes the characteristics of these included studies.

**Table 3.**
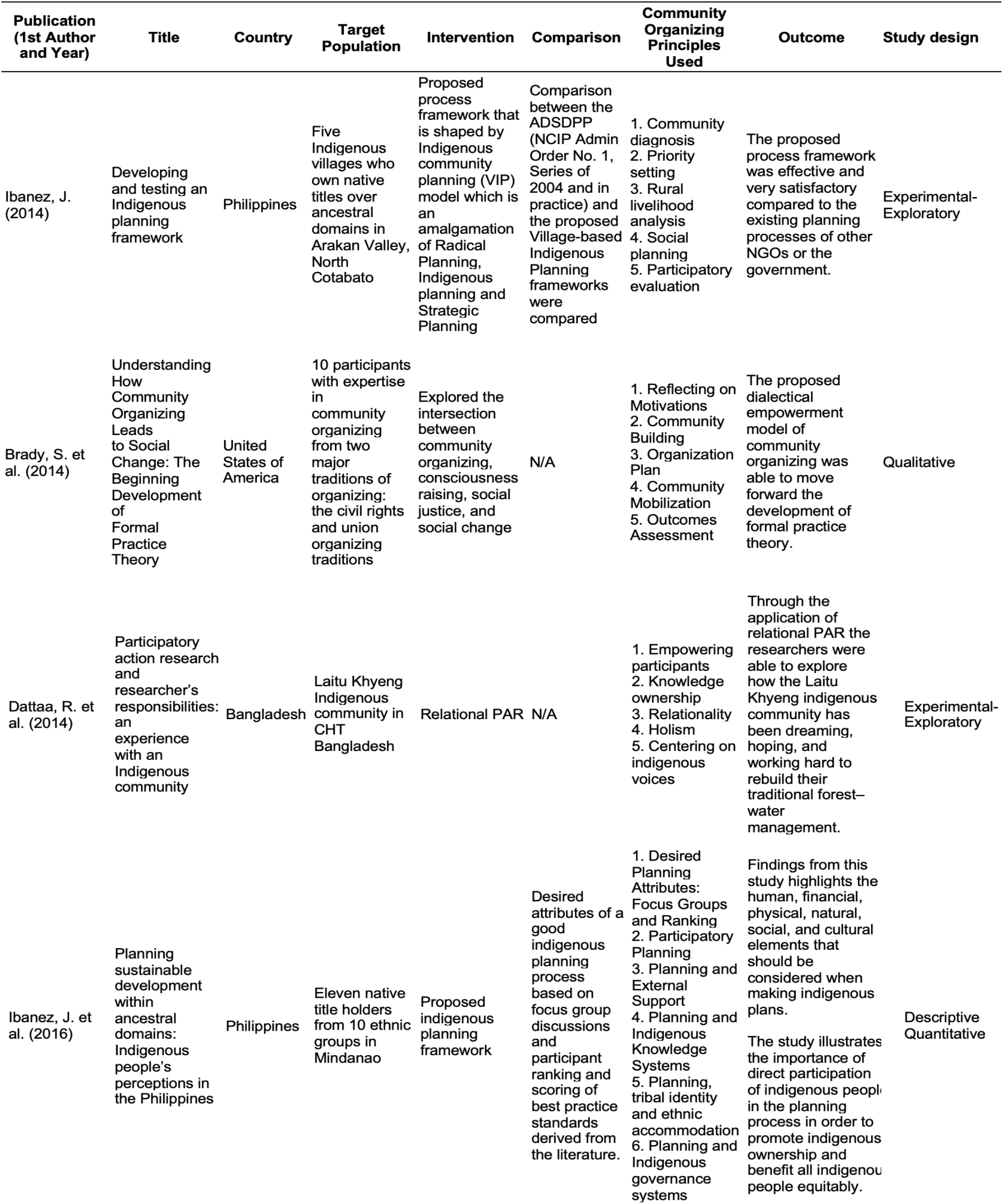
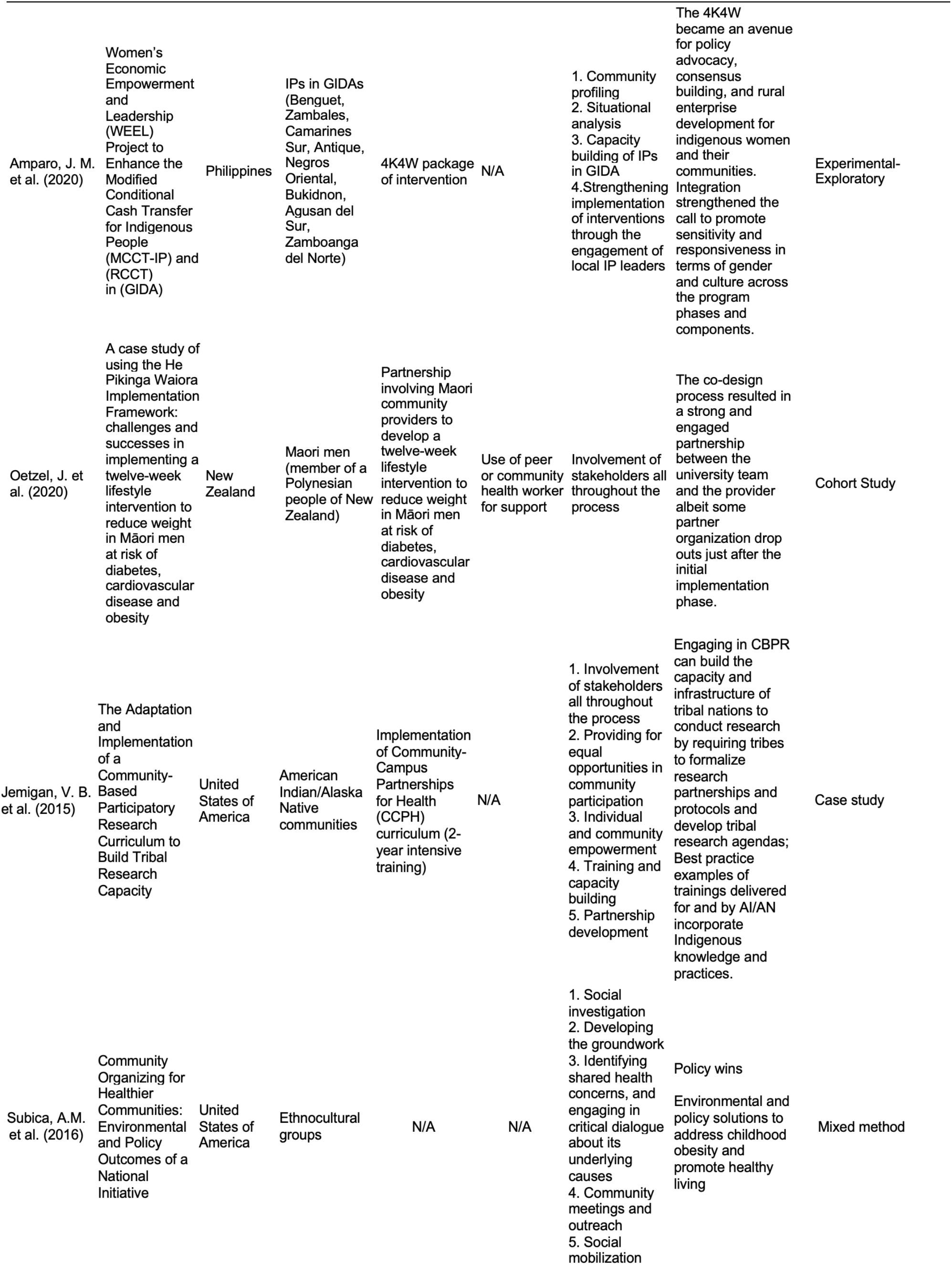
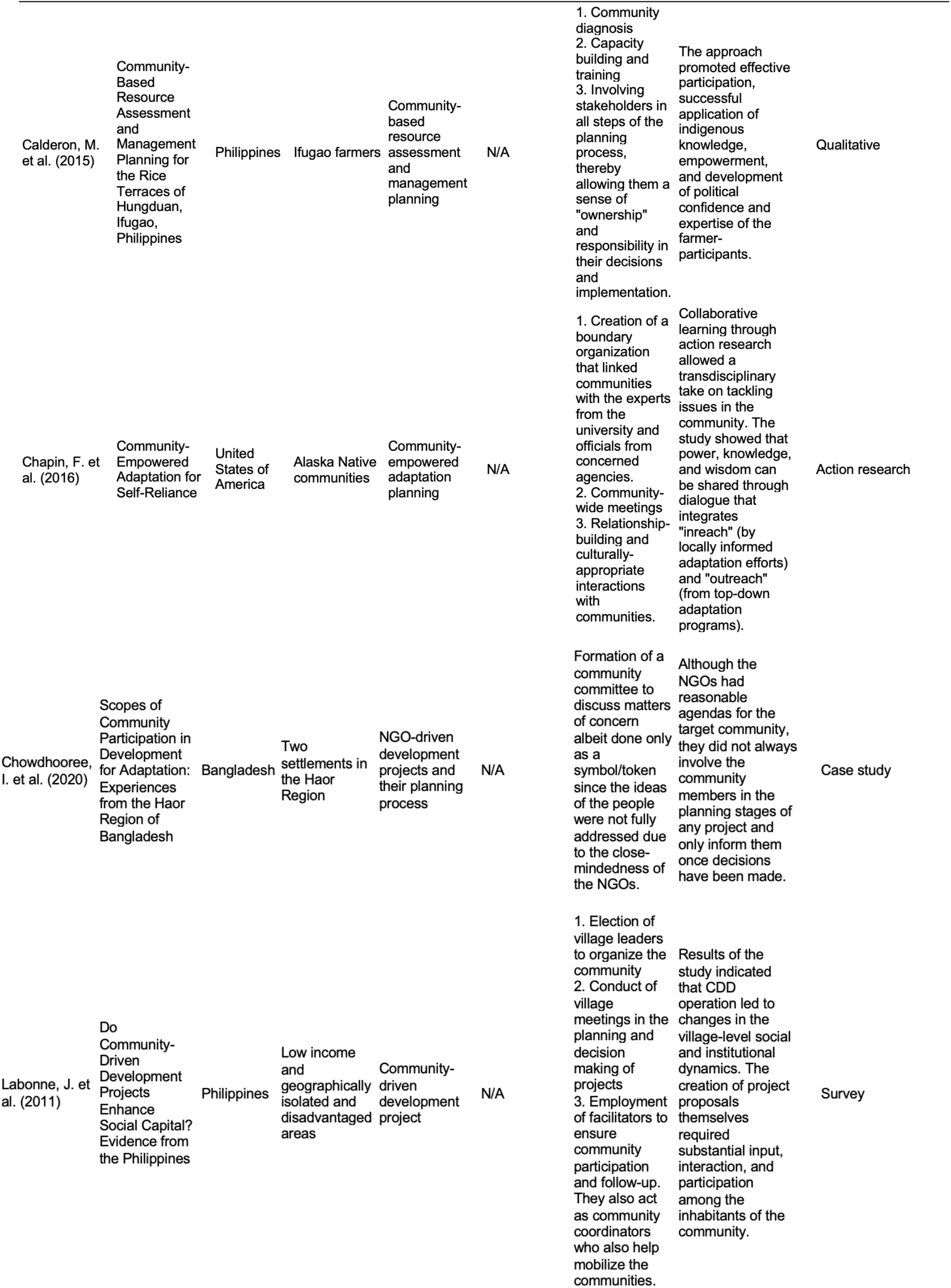
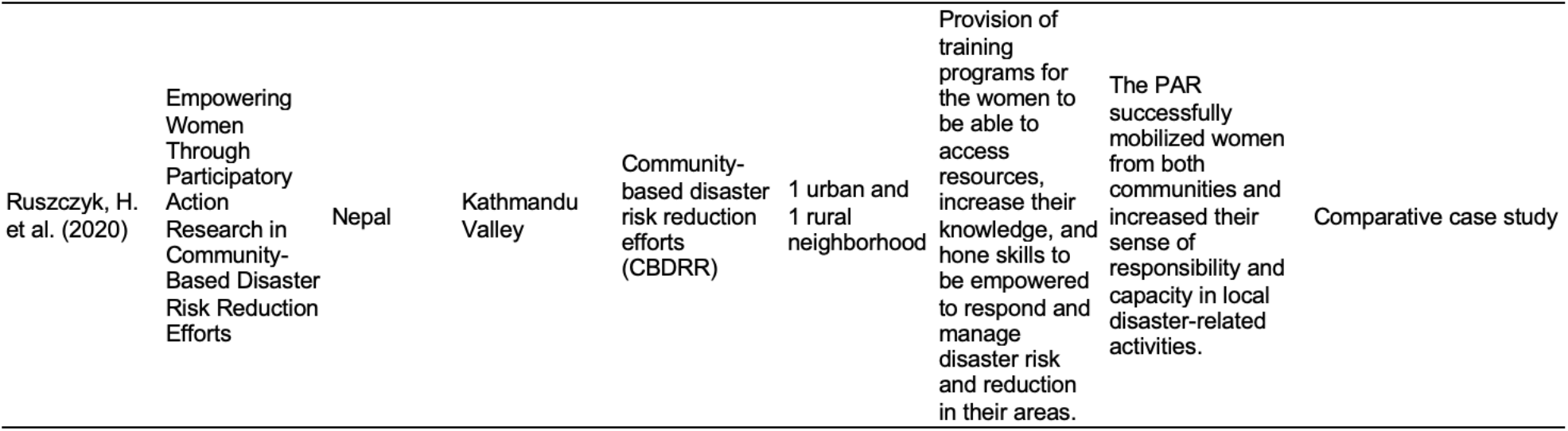
Characteristics of the Included Studies

## 4.0 DISCUSSION AND IMPLICATIONS

### 4.1 Proposed Community Organizing Approach for Indigenous People in the Philippines

In this systematic review, the researchers were able to identify community organizing principles used in programs directed for the indigenous population. The researchers categorized the different principles to the following stages: Pre-entry, Entry, Organization Building, Strengthening, and Turn Over Phase. The researchers proposed a community organizing framework from the themes and best practices that surfaced during the review. Methodologies and evaluation plans for each phase of the community organizing process were also identified. Building on knowledge presented, this paper contributes to the gaps in existing framework used for community organizing among the IP population in the country. Figure 3 presents the proposed Community Organizing Protocol for Indigenous People in the Philippines.

**Figure 3.**
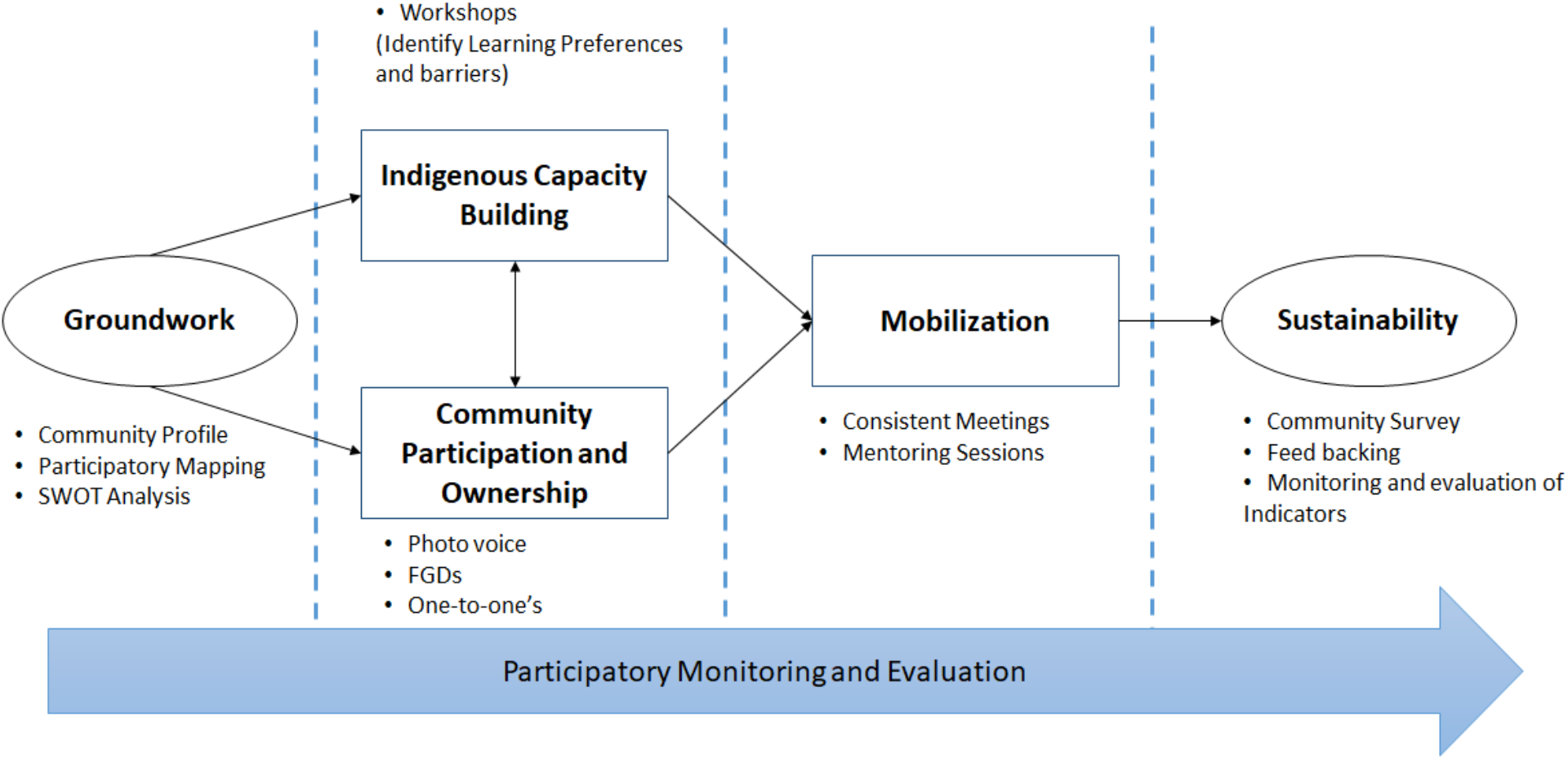
Proposed Community Organizing Protocol for Indigenous People in the Philippines.

#### Groundwork

The groundwork is the preparatory phase for community organizing. This constitutes pre-work activities that set the foundation of reform strategies before engaging with the community. The reviewed studies documented groundwork activities such as conduct of community diagnosis, community profiling, social investigation to describe the system. The study of Oetzel J. et.al (2020), and Jemingan V. et. al (2015) also suggested the involvement of stakeholders and research partners such as the academe in the community organizing process.

Groundwork activities are critical because it examines the local context and conditions in the village using the subjects: people, ancestral domain, and resources^23^. Some literature also stated that learning the local culture and governance structures as well as identifying respected leaders and key decision makers who allocate resources are also essential before entering the community.^24^

Indigenous leadership structures should be identified. The acceptance and involvement of local leaders and potential leaders as well as knowing how to deal with them are critical in mobilizing the community. Leaders should also be approached respectfully in accordance with local cultural practices.^24^

The paper also suggests that there should not be a pan-indigenous understanding. There must be a full recognition of the diversity of the indigenous people and understanding of local history, cultures, powers and resources^31^.

Community organizers can make use of secondary data sources as well as Participatory Research Appraisal (PRA) tools to examine the setting and power levels within the community^23^.

#### Indigenous Capacity Building

Indigenous capacity building is necessary to prepare the community members. Organizers must ensure that the community has the required skills set and be provided with mentoring and coaching sessions to guide them in the process. This paper was able to identify activities such as Community Building, Empowering Participants, and Training as capacity building activities in the community organizing process.

Provision of skills training and inputs to indigenous communities may pose challenges. A study conducted in Canada identified that indigenous people have learning preferences such as being visual, spatial, and being comfortable in a small learning group. Some learning barriers identified are namely language, and learning difficulties due to illiteracy^32^.

The researchers categorized this step as part of the cognitive intervention in the community organizing process. This is tailored with a behavior-centered intervention specifically in increasing the ownership of the community members.

#### Community Participation and Ownership

To address the root causes of health and social problems in the community requires a deep partnership within the community members. Meaningful engagement can take place through members of the community understanding their personal role and counterpart in the development process and success of the program. However, participation is not enough to ensure complete community engagement, a shared vision is required to build commitment.

Grassroots community organizing groups may build interpersonal relationships among participants through awareness-raising activities such as photovoice, focus group discussions, semi structured conversations between two people also known as one-to-ones. Planning and forming of action plans should be done with the community is instrumental in ensuring that integration of indigenous knowledge aligns with culturally safe intentions, and to avoid the appropriation of knowledge^31^.

#### Mobilization

After building the capacity of the core group, this phase entails the actual implementation. Based on the reviewed studies, community facilitators or coordinators play an important role in ensuring the success of community participation during project implementation by helping in mobilizing communities as well as in ensuring adequate representation^33,34^. The facilitators motivate the community to willingly agree in the fulfillment of the project and also encourage the community to participate in decision making.

Moreover, consistent meetings were found to be beneficial in ensuring proper management and execution of a project/program/ intervention in the community^34^. Project planners were able to closely monitor the implementation during site visits. These frequent visits also helped in increasing participation among residents in the community. Regular community meetings may also promote reciprocal learning and establish trust and respect^24^. Aside from the consistent meetings, open communication with project planners also aid in the success of organizing indigenous communities. Transparent communication with and within the community (e.g. community-wide meetings) may increase the likelihood that a broad spectrum of issues and views would be discussed^35^. This is also one of the steps included in the eight-step approach aligned with the CARE model which is to facilitate continued, multimethod communication. Interpersonal communication is often the most important means of information sharing at the community level^24^.

A study conducted in Bangladesh also presented various methods used in engaging community participation in indigenous communities that were aligned with the Participatory Action Research (PAR). Data analysis was emphasized as one of the significant parts of the study (aside from sharing research results), wherein the elders and knowledge holders also took part in the sharing of codes in which their core values, beliefs, and spiritual practices were also considered in the process^36^.

Public health practitioners can use these tools and methodologies to ensure success in their community organizing efforts.

#### Sustainability

This phase focused on maintaining the coalition, as well as the implemented programs or projects in the community. Based on the reviewed studies, stakeholders should be involved all throughout the process so that they may feel a sense of “ownership” and are accountable for the success of the program/ intervention ^37^. One of the critical factors identified in the success of indigenous community-managed programs is through facilitating community ownership and control. This can be embedded in the community-managed programs by forming local indigenous management bodies and formal agreement with partner organizations.^25^

In another study reviewed, ‘sharing data analysis’ was also found to be useful in maintaining programs in the community^36^. Dissemination of the findings to the key stakeholders is necessary for them to understand the context and thereby sustain its implementation^38^. One of the community organizing principles involves consciousness-raising through experiential learning^16^. Data sharing is a means of increasing awareness and motivation among people for them to sustain what has been introduced to the community.

In addition, monitoring and evaluation is also essential in the continuity of the programs/projects. Tracking progress, successes, failures, and costs are also critical for sustainability^24^. Results should also be regularly communicated to the beneficiaries and partners so that these will be incorporated in the strategic action planning with the community^39^. Thus, there is a need to establish indicators and a monitoring scheme ^38,23^. Moreover, formation of a core working group is also needed in this phase since they are able to conduct the evaluation and therefore, help in ensuring its continuity ^34,35^.

#### Participatory Monitoring and Evaluation

Among the articles reviewed, only the study conducted by Ibanez J. (2014) explicitly stated participatory evaluation. Brady et.al (2014), suggested Outcomes assessment. The researchers identify the inclusion of an evaluation protocol as a vital component in the community organizing framework. The proposed framework makes use of the Participatory Monitoring and Evaluation (PM&E) as the monitoring and evaluation approach. PM&E is defined as applied social research that involves a partnership between trained evaluation personnel and practice-based decision makers, organizational members with program responsibility, or people with a vital interest in the program^40^. This approach is essential for evaluating community organizing programs as this build on the involvement of the community in every step of the process. A comprehensive monitoring and evaluation system should enable the organizers, community members, and leaders to learn which strategies work and what needs to be improved.

The PM&E process will be able to cover all phases of the organizing process. This includes 1. Participatory Appraisal, 2. Participatory Planning and Project Design 3. Participatory Development of Baseline Indicators 4. Participatory Baseline Data Collection 5. Participatory Monitoring and Evaluation Plan Design 6. Participatory Implementation 7. Participatory Monitoring and Review 8. Participatory Evaluation 9. Feedback and Participatory Decision Making^41^. All steps are done in consultation and collaboration with organizers, funders and the community beneficiaries, deciding what will be monitored and how the monitoring will be conducted. Together, they analyze the data gathered through monitoring and assess whether the project is on track in achieving its objectives. Participatory monitoring enables project participants to generate, analyze, and use information for their day-to-day decision making as well as for long-term planning.

### 4.2 Limitations

Some relevant studies might have been missed in the review because they were published in languages other than English. There was also no funding utilized thus, the researchers were limited to searching open-access scientific databases which may not capture other peer-reviewed journals. No primary data gathering and no face-to-face interview were done during the course of the research. All methods, techniques, and communication were done online by the researchers.

The community organizing approach presented in the review are best recommended practices, nevertheless this should still be reviewed upon by the National Commission on Indigenous People in order to verify that every phase is in the light of the IPRA. Though most of the frameworks used by the articles were applied in a study population, the proposed community organizing approach have yet to be tested in indigenous communities.

### 4.3 Conclusion

The shortcomings of the existing framework used in the Philippines by the government as discussed in the earlier sections can be minimized or supplemented by the proposed community organizing protocols. The Indigenous Planning Framework has a wide range of scope and uses a needs-based approach wherein it mainly focuses on the deficit of the community. Although the IP’s concerns are considered in searching for promising opportunities, the problem-based approach can overemphasize what the community lacks in order to attract outside aid. This may affect program implementation and sustainability especially if the problems are already addressed or the organizers have already achieved their goals. Which is why the researchers have recommended an asset-based approach in community organizing protocols in order to empower the community through collective engagement and have a control over their own resources.

The framework was divided into five domains: Groundwork, Indigenous Capacity Building, Community Participation and Ownership, Mobilization, and Sustainability. Groundwork is a very essential process in Indigenous communities and was incorporated in the Indigenous People Framework as a Situational Analysis step. Various methods and tools such as Stakeholder analysis were suggested in the existing IP Framework in order to have an inclusive and responsive project. In this study, there are a wide range of tools identified in the Groundwork phase that have been proven effective in analyzing the conditions of Indigenous communities. This phase analyzes the different social and cultural structures and understands the land and natural resources that are linked to their identities and livelihoods which is essential in establishing rapport in the IP community.

The study also highlighted the need to focus on the first two phases of the community organizing process which is Community Participation and Capacity Building in order for the intervention to be socially acceptable. Since the IP communities are governed with their own indigenous institutions, appropriate consent is necessary before any activities can be taken within their ancestral domains. The integration of the findings obtained in the first two phases within the indigenous structure of social preparation activities are important as an entry point for community organizing in IPs. It is important to work within the social structures and system thus; the community should have a full understanding of the project to increase acceptance and support of the community organizing process. On the other hand, methods such as focus group discussions, photovoice, and common place books are uniquely designed for IPs so as to promote engagement while overcoming the language barriers among the community. Participation of indigenous groups is integrated in all phases of community organizing in order to establish a strong sense of ownership of the project. Monitoring and Evaluation should be observed in all phases and be conducted in a participatory manner in order to ensure that the steps are responsive to the needs of the community and promote the value of their cultural heritage.

Employing a holistic community organizing strategy for IPs will facilitate the development of a community-based system that utilizes maximum participation and cultural empowerment at the same time sustaining cultural integrity. The study suggests the best and effective practices and tools that can be used in order to have meaningful participation towards IPs. Needless to say, interventions should be tailored and best developed on a community level basis by stakeholders and community organizers to ensure all factors will be considered.

### 4.4 Future Research

In order to evaluate the effectiveness of the proposed community organizing protocol, testing the framework on indigenous communities in the Philippines is recommended. The proposed approach is not being suggested as a cure-all formula nor as a replacement of the existing community organizing process, however it is recommended to apply it on existing programs of the government or NGOs in order to assess its effectiveness. Due to the complexity of community organizing in indigenous communities, this should be verified on a community-level basis by the IP organizers and researchers working closely with the community. Needs analysis and primary data gathering is highly recommended to know the strength and weaknesses of each tool since the findings on the general population do not always accurately reflect indigenous communities due to their distinct social patterns. The proposed protocol should be undertaken in close coordination with the National Commission on Indigenous People at the national and local levels, while at the same time ensuring that the four bundles of rights defined by IPRA are met (Right to Ancestral Domains and Lands, Right to Self-Governance and Empowerment, Right to Social Justice and Human Rights, and Right to Cultural Integrity).

## Data Availability

The authors confirm that the data supporting the findings of this study are available within the article [and/or] its supplementary material.

